# The association between influenza vaccination, all-cause mortality and cardiovascular mortality: a protocol for a living systematic review and prospective meta-analysis

**DOI:** 10.1101/2021.08.31.21262935

**Authors:** Rong Liu, Anushka Patel, Xin Du, Hueiming Liu, Bette Liu, Chi Wang, Gian Luca Di Tanna

## Abstract

**Introduction:** Influenza virus infection is known to increase the risk of cardiovascular events, especially in populations with pre-existing cardiovascular disease. Considering that influenza is vaccine preventable, international guidelines recommend high-risk populations with CVD receive an influenza vaccine every year, but there are various classifications of recommendations and levels of evidence. Previous systematic reviews concluded uncertain evidence on influenza vaccine efficacy for preventing cardiovascular events in the general population or in populations with pre-existing CVD. Limited safety data of influenza vaccines were reported for populations with pre-existing CVD. Randomized control trials with larger sample sizes relative to previous studies are emerging, the findings of these trials are likely to be highly influential on summary efficacy estimates.

**Methods and analysis:** We aim to perform a living systematic review and a prospective meta-analysis to evaluate the efficacy and safety of influenza vaccines compared to no vaccines or placebo for preventing mortality or cardiovascular disease events in the general population and in populations with pre-existing CVD.

**Ethics and dissemination:** Formal ethical review is not required as this study does not need primary data collection. We will publish results of the living systematic review and prospective meta-analysis in a peer-reviewed journal. Findings will also be presented at relevant meetings.

**PROSPERO registration number:** CRD42021222519.

**Strengths and limitations of this study:**

- The living systematic review will continually incorporate the latest research findings and keep the synthesized information updated. A prospective meta-analysis will better address this evolving evidence.
- Safety of influenza vaccines in populations with pre-existing cardiovascular diseases will be studied in particular to complete the current evidence base.
- Observational studies may affect the overall quality of the study results. We will stratify the analysis by study design and present both randomized and non-randomized results.

## Introduction

Influenza virus infection is known to increase the risk of cardiovascular events, especially in populations with pre-existing cardiovascular disease. ^1^ The World Health Organization (WHO) recommends countries aim for 70% influenza vaccine coverage for high-risk groups, including the elderly and individuals with known chronic conditions. ^2^

Cardiovascular disease (CVD) takes approximately 18 million lives each year, which accounts for one third of all deaths worldwide. In order to achieve the global target of “25 by 25” and “1/3 by 30”, reducing a quarter of premature deaths from NCD by 2025 and one-third of them by 2030, effective interventions need to be identified and implemented in the most vulnerable populations. ^3^ The economic burden of CVD is projected to be more than one trillion dollars in 2030, half of which relates to direct medical costs. Cost-effective interventions are needed to flatten the rising curve of healthcare costs for CVD. ^4,5^ A modeling study showed a fully funded influenza vaccination program compared to a self-paid one was cost-effective in population over 60 years old in China, with vaccination coverage rate being 30% versus 0% respectively. An influenza vaccine coverage of 30% would avert 8,800 influenza-associated excess death attributable to respiratory causes per year, which accounted for 98% of all costs from outpatient consultation, hospitalization, death, and loss of productivity. ^6^

Considering that influenza is vaccine preventable, international guidelines recommend high-risk populations with CVD receive an influenza vaccine every year, but there are various classifications of recommendations and levels of evidence. ^7,8^ This uncertainty is reflected in the most recent 2015 Cochrane systematic review, which concluded uncertain evidence on influenza vaccine efficacy for preventing cardiovascular events in the general population or in populations with pre-existing CVD. ^9^ The uncertainty origins from risks of bias in pooled studies and higher-quality evidence is in need to confirm the findings.

This review included 8 randomized controlled trials (RCTs) of influenza vaccine versus placebo or no vaccine with a total of 12,029 participants, and searched literature between the starting dates of database archive and October 2013. Their meta-analysis, pooling 4 of the included trials which assessed the association between influenza vaccination and cardiovascular mortality, showed a pooled risk ratio (RR) of 0.45 (95% Confidence Interval (CI) 0.26-0.76 [p=0.003]). With more RCTs and a number of large-scale observational studies having also been conducted since the Cochrane review, it is useful to update our understanding of the current evidence on influenza vaccines for preventing CVD events in both the general population and high-risk groups. ^10-13^

One more recent systematic review and meta-analysis has been conducted, pooling four RCTs (n=1667) and twelve observational studies (n=235 391) and indicating a pooled RR of 0.87 (95% CI, 0.80–0.94 [p<0.001]) for major adverse cardiovascular events among patients receiving influenza vaccines versus those receiving no vaccine or a placebo. ^14^ This systematic review focused on the use of influenza vaccine as a secondary prevention measure for patients with established CVD and extracted articles published through to January 2020.

Two trials RCT-IVVE (NCT02762851) and IAMI (NCT02831608) comparing inactivated influenza vaccine to placebo in patients with either heart failure or recent acute myocardial infarction are still ongoing. ^12,13^ These trials, specifically focusing on myocardial infarction (MI) (n = 4,400) or heart failure (HF) (n = 5,000), respectively, are expected to report results in 2021 or later. ^12,13,15^ With their large sample sizes relative to previous studies, the findings of these trials are likely to be highly influential on summary efficacy estimates.

While the safety of influenza vaccines in the general population is well established, there is a paucity of safety data in populations with existing chronic disease. In these populations, current synthesized data on adverse event following immunization (AEFI), or adverse health events or health problems that may be vaccine-attributable are sparse.

Perhaps as a consequence of persistent uncertainty relating to both efficacy and safety, influenza vaccine coverage rates (VCR) are variable and often low in populations with pre-existing CVD. For example, recent influenza VCR in heart failure patients ranges from nearly 0% in Asia to approximately 80% in Europe. ^16^ In particular, most low- and middle-income countries (LMIC) have not reached the target of 70% VCR set by WHO for high-risk groups. In China, the estimated influenza VCR for the entire population is 2% and is even lower (<1%) among high risk groups. ^16,17^ In a limited number of Chinese cities with a policy for free influenza vaccination among seniors, VCR in those older than 65 years is reported to be around 20%. ^18^

As evidence from ongoing RCTs are still emerging, it is appropriate to conduct a living systematic review (LSR), which will continually incorporate the latest research findings and keep the synthesized information updated. ^19^ Along with the LSR design, a prospective meta-analysis (PMA) will better address this evolving evidence. Through a PMA, we will aim to include studies that currently are still not published to avoid potential bias. ^20^

## Methods

This protocol follows the 2015 Preferred Reporting Items for Systematic Review and Meta-Analysis Protocols (PRISMA-P) statement, along with the elaboration and explanation report, and the checklist. ^21,22^

### Objective

The overall objective of this LSR is to evaluate the efficacy and safety of influenza vaccines compared to no vaccines or placebo for preventing mortality or cardiovascular disease events in the general population and in populations with pre-existing CVD.

### Eligibility criteria

Studies selected for inclusion will meet the following Population, Intervention, Comparator, Outcome, and Study design (PICOS) criteria.

#### Population

We will include studies focusing on the general population aged 18 years and above, or populations with an established history of CVD. CVD is defined to include any diagnosis of hypertension (high blood pressure), coronary heart disease (heart attack), cerebrovascular disease (stroke), peripheral vascular disease, heart failure, rheumatic heart disease, congenital heart disease, or cardiomyopathies.

#### Intervention

We will include studies that investigate the effects of inactivated influenza vaccine or live attenuated influenza vaccines during any influenza season, regardless of the valency, dose, administration route, boosts and use of concomitant vaccination strategies.

#### Comparator

We will include studies of no vaccine or placebo as comparators.

#### Outcomes

Outcomes of interest are all-cause mortality, cardiovascular-specific mortality, all-cause hospitalization, and cardiovascular-specific hospitalization or events. Cardiovascular events include any diagnoses due to myocardial infarction, heart failure, or stroke.

#### Types of studies

We will include any type of RCT (Individually randomized, cluster, stepped wedge, other) and observational study (cohort and case-control). We will include published or accepted articles with RCT or observational designs without date limits. Pre-prints, theses, or dissertations without formal peer-review will not be included. No language restrictions will be imposed on the search strategies. We will include studies conducted in hospital-based, community-based, or long-term care facility-based settings.

#### Time frame

We will include studies reporting outcomes with the follow up period not surpassing the starting point of the next influenza season. The living status of the systematic review will be maintained for a minimum of three years after protocol publication. The baseline living systematic review and prospective meta-analysis are planned to start from June 2021. An update will be performed every 6 months after the baseline.

### Information sources

We will search the following databases:

- Cochrane CENTRAL; ^23^
- ClinicalTrials.gov; ^24^
- The Chinese Clinical Trial Registry (ChiCTR); ^25^
- Medline (PubMed interface);
- Embase (Ovid interface);
- CNKI; ^26^
- Wanfang; ^27^
- The Database of Abstracts of Reviews of Effects (DARE); ^28^
- The Economic Evaluation Database (EED); ^29^
- Health Technology Assessment (HTA). ^30^

Each database will be searched separately by two authors with an initial search strategy developed from PubMed and then adapted for other databases. We plan to search the reference lists of eligible articles and contact corresponding authors of papers for missing information.

### Search strategy

We will use the search strategy of the previous Cochrane review by Clar et al. (**Appendix 1 and 2**). ^9^ Keywords of ‘Influenza Vaccines’ and ‘Cardiovascular Diseases’ will be used to capture observational studies. Auto alerts will be configured to receive monthly updates.

### Study records

#### Data management

The search results from all databases will be imported into the reference management software EndNote X9. Duplicated reports from the same study will be removed. The unique records will be imported to the study screening and data extraction software Covidence. ^31^

#### Selection process

Predefined inclusion and exclusion criteria will be used for screening. After title and abstract screening, full texts will be downloaded for the remaining studies. Study tags will be created to mark predefined eligibility criteria for easier screening and post-hoc checks. The entire selection process will be conducted independently by two reviewers. Conflicts or disagreements between the two reviewers will be resolved by a third reviewer. A screening process flowchart will be presented as per PRISMA recommendation.

LSR specific indicators will also be reported. These will include, for example, LSR version number, time since preceding update, number of citations screened for the LSR update period, number of identified newly published eligible primary study protocols, number of identified newly published eligible primary studies, disposition of newly identified eligible primary studies (i.e., incorporated or not). Changes in LSR methodology compared to previous versions will be reported. Any changes in statistical results, certainty of evidence, and conclusions from previous iterations will be highlighted in the LSR report. Differences between the protocol and the review will be recorded and justified.

#### Data collection process

Data will be extracted and entered into a predefined data extraction form. Data extraction will be done by two authors independently, with discrepancies resolved by a third author. The data extraction form will be reviewed by the entire review team and piloted for the first three studies before its roll out.

### Data items

The following data elements will be extracted:

- General information: title, authors, author contact details, year of publication, journal, language, type of paper (original research, protocol, review, and editorial).
- Population: inclusion and exclusion criteria, sampling and recruitment methods, study population characteristics and comparability between groups at baseline (age, sex, socio-economic status, country, inpatient or outpatient, comorbidity, and concomitant treatment regimen other than vaccination). General population or population with pre-existing CVD, and the disease subtype (MI, HF, stroke etc.) if with pre-existing CVD. Coronavirus disease (COVID) status.
- Intervention: vaccine type, valency, dose, administration route, timing of vaccination, number of participants in intervention group, duration of follow up. Level of the match of influenza vaccines to circulating strains.
- Comparator: placebo, or no vaccine, number of participants in control group, overall follow up, duration of follow up.
- Outcomes: definition, time points measured, number of outcome events in intervention and control group, incidence rate in intervention and control group, prevalence in intervention and control group, unadjusted and adjusted effect measures (odds ratio [OR], risk ratio [RR], or hazard ratio [HR]), covariates used for adjustment, effect size (point estimate, standard error or standard deviation or confidence interval [CI]), missing data, reason for missingness, approach to handling missing data, statistical methods, randomization process. Dropout rate, loss to follow-up rate, and adverse event rate in intervention and control groups.
- Study design and methods: study type, registration number, country and setting, recruitment time, date of first participant (or cluster) randomized, date of last participant (or cluster) randomized, date of last participant followed up for outcomes in RCTs, date of first participant recruitment, date of last participant followed up for outcomes in observational studies, vaccination date, hemisphere, match of the influenza vaccine strains to those circulating, reporting time, study duration, study objectives.
- Study funding and conflict of interest.

Effect sizes will be extracted as reported in the source article, and transformed when appropriate. In case of missing information from an included paper, an attempt of contacting the authors to obtain these data will be made.

### Assessment of risk of bias in individual studies

We will apply Version 2 of Cochrane risk-of-bias tool (RoB 2) to included RCTs. ^32,33^ Through RoB 2, studies will be assessed across a number of domains, including random allocation sequence, allocation sequence concealment, blinding, outcome assessment, missing data, analysis methods, to classify studies into a ‘low risk of bias’, ‘some concerns of risk’, or ‘high risk of bias’ categories.

For observational studies, we will apply the Risk Of Bias In Non-randomized Studies – of Interventions (ROBINS-I). ^34^ ROBINS-I assesses a number of domains, including confounding, selection bias, baseline comparability between groups, intervention fidelity, outcome measurement, selection of reported results. Studies will be classified into categories reflecting risks of ‘low’, ‘moderate’, ‘serious’ bias. Two authors will perform the RoB assessment with consensus.

### Data synthesis

#### Narrative synthesis

A narrative summary of the effect of influenza vaccines on outcomes will be provided. Study characteristics (design, participants, intervention, comparator, outcomes, methods, funding, and conflict of interest) will be presented in a table.

#### Criteria for quantitative data synthesis

We will perform separate meta analyses pooling results from observational studies and RCTs and carefully evaluate differences between the two sets of results as we expect observational studies to report higher effects than the ones from trials. We plan to carry out a Hartung-Knapp-Sidik-Jonkman random effects meta-analysis whenever it will be feasible to do so. If it is not feasible, we will revert to narrative review (for instance if only 1 study reported a specific outcome). We will also report pooled effects according to the common (i.e. fixed) effect model. To better handle the uncertainty of the various parameters to be estimated (especially the between-study variance) and also considering the prospective nature of this study, we will also perform a Bayesian Meta Analysis: this will allow us to calculate probabilities of the vaccine to be effective and also, as data accrue, the posterior distribution of the pooled effect will be updated to reflect information derived until that moment that will be used as an informative prior distribution of the effect size. Sensitivity analyses will be performed using vague and vaguely informative priors for the effect size and for the between-study heterogeneity.

For binary outcomes, we will use risk ratio (RR) with 95% confidence interval (CI) (or credible intervals, for the Bayesian Meta-Analysis) to measure the effect of influenza vaccination. For time-to-event data, we will use hazard ratio (HR) with 95% CI accordingly.

#### Unit of analysis issues

Analyses will be done at a study level. For cluster randomized trials (including stepped wedge trials, if any) we will ensure the cluster effect has been taken into account. If not, we will inflate standard errors using the design effect (which is a function of the average cluster size and intra-class correlation coefficient). If the intra-class correlation (ICC) coefficient is not reported, we could “borrow” the ICC from one study and apply to the other CRTs if missing, or run sensitivity analyses by various design effect inflation factors.

#### Dealing with missing data

For studies without reported data for an outcome of interest, we will try to obtain this information by contacting the original authors. The impact of excluding studies with high level of missingness will be explored in sensitivity analyses.

#### Assessment of heterogeneity

We plan to assess heterogeneity by formal test of homogeneity and evaluating the proportion of variability attributable to heterogeneity rather than sampling error using the *I*^2^ statistic. As per the Cochrane Handbook we will consider values of *I*^2^ between 50% and 90% as substantial heterogeneity and above 75% as considerable heterogeneity. Subgroup analyses and meta regression based on following variables will be used to explore possible reasons of heterogeneity:

- Population type (general population, pre-existing CVD population, COVID population);
- Age groups (if feasible);
- Hospitalized versus outpatient;
- If disease-specific population, then consider the severity of disease, for example, ejection fraction category for patients with heart failure;
- Non-pandemic years versus 2009/2010 pandemic year if feasible;
- Level of the match of influenza vaccines to circulating strains if reported;
- Level of risk of bias.

We plan to investigate the likelihood of selective outcome reporting bias by comparing the study report and its corresponding protocol. ^35^ If more than 10 studies are finally selected, formal Egger’s regression-based test and eye-ball assessment of the funnel plots will be explored to evaluate small-study effects.

### Confidence in cumulative evidence

We will use the Grading of Recommendations Assessment, Development and Evaluation (GRADE) system to judge the overall quality of all findings. ^36^ GRADE system classifies evidence into ‘high quality’, ‘moderate quality’, ‘low quality’, and ‘very low quality’, based on methodology quality, consistency, directness, precision, and the risk of reporting bias. The cumulative quality of evidence will be assessed by all authors.

### Patient and public involvement

No patient involved.

## Discussion

### Main findings of previous reviews

The 2015 Cochrane review included four trials (n = 1,682) of influenza vaccination compared with placebo or no vaccination for preventing cardiovascular mortality in populations with pre-existing CVD. It presented a wide CI of the pooled RR for preventing cardiovascular mortality by influenza vaccines (0.45, 95% CI: 0.26 to 0.76, *P* = 0.003). The pooled studies had some risk of bias and was of small sample size, which may lead to the wide CI of estimated efficacy. Not enough evidence was available to establish whether influenza vaccination has a role to play in the primary prevention of cardiovascular disease. With ongoing trials aiming to recruit more than 9,000 participants, pooling these results together would help strengthen evidence base.

### Impact and significance of the review

Although current guidelines recommend populations with pre-existing CVD receive annual influenza vaccinations, there is inconclusive evidence regarding the efficacy and safety of influenza vaccines for preventing death or hospitalization from cardiovascular disease. ^7,8^ Previous systematic reviews reached uncertain conclusions with a lack of high-quality studies. As several large ongoing trials are investigating influenza vaccine for preventing cardiovascular events, new evidence is accumulating and may substantially add to the evidence base. This provides an important opportunity to update current literature on the efficacy/effectiveness of influenza vaccine on cardiovascular mortality and hospitalization.

The living systematic review will continuously synthesize the latest research findings so as to inform the public and healthcare professionals. With the most updated evidence regarding the efficacy of influenza vaccination in preventing CVD morbidity and mortality especially in high-risk populations, health care providers may be able to make recommendations to individual patients with more certainty. From a public health point of view, the findings may influence vaccine policies in relation to the general and high-risk populations.

This review will also provide important pooled parameters estimates that can also inform subsequent economic evaluations that will assess cost effectiveness profiles of various vaccines and vaccination strategies.

### Registration

This LSR protocol will be registered with the International Prospective Register of Systematic Reviews (PROSPERO). ^37^ The registration ID number is CRD42021222519.

### Editorial and publication process consideration

LSR versions will be submitted to a peer-reviewed journal accommodating iterative versions of the same systematic review.

## Supporting information

PRISMA-P Checklist

Search Strategy

## Data Availability

Aggregate data will be extracted from published papers

## Author statement

All authors have contributed to the final submitted manuscript. Gian Luca Di Tanna is the lead supervisor of the first author. Anushka Patel, Xin Du, and Hueiming Liu are co-supervisors. Rong Liu drafted the initial manuscript and received guidance regarding the content area and methodology from all authors. Chi Wang is the second reviewer. All authors have read and agreed the final manuscript.

## Data statement

The search strategy is in the appendix online. Data extraction form is accessible upon reasonable request.

