## Supplementary material for "The association between influenza vaccination, all-cause mortality and cardiovascular mortality: a protocol for a living systematic review and prospective meta-analysis": Search Strategy

### Appendix 1. Search strategies 2013 - cardiovascular disease

#### The Cochrane Library

#1MeSH descriptor: [Influenza Vaccines] this term only
#2(influenza* near vaccin*)
#3(influenza* near immuni*)
#4(flu near immuni*)
#5(flu near vaccin*)
#6(#1 or #2 or #3 or #4 or #5)
#7MeSH descriptor: [Cardiovascular Diseases] explode all trees
#8cardio*
#9cardia*
#10heart*
#11coronary*
#12angina*
#13ventric*
#14myocard*
#15pericard*
#16isch?em*
#17emboli*
#18arrhythmi*
#19thrombo*
#20atrial next fibrillat*
#21tachycardi*
#22endocardi*
#23(sick next sinus)
#24MeSH descriptor: [Stroke] explode all trees
#25(stroke or stokes)
#26cerebrovasc*
#27cerebral next vascular
#28apoplexy
#29(brain near/2 accident*)
#30((brain* or cerebral or lacunar) near/2 infarct*)
#31MeSH descriptor: [Hypertension] explode all trees
#32hypertensi*
#33(peripheral next arter* next disease*)
#34((high or increased or elevated) near/2 blood pressure)
#35MeSH descriptor: [Hyperlipidemias] explode all trees
#36hyperlipid*
#37hyperlip?emia*
#38hypercholesterol*
#39hypercholester?emia*
#40hyperlipoprotein?emia*
#41hypertriglycerid?emia*
#42MeSH descriptor: [Arteriosclerosis] explode all trees

#43MeSH descriptor: [Cholesterol] explode all trees
#44cholesterol
#45"coronary risk factor*"
#46MeSH descriptor: [Blood Pressure] this term only
#47"blood pressure"
#48#7 or #8 or #9 or #10 or #11 or #12 or #13 or #14 or #15 or #16 or #17 or #18 or #19 or #20 or #21 or #22 or #23 or #24 or #25 or #26 or #27 or #28 or #29 or #30 or #31 or #32 or #33 or #34 or #35 or #36 or #37 or #38 or #39 or #40 or #41 or #42 or #43 or #44 or #45 or #46 or #47
#49#6 and #48

**MEDLINE Ovid**

1. Influenza Vaccines/
2. (influenza$ adj3 immuni$).tw.
3. (flu adj3 vaccin$).tw.
4. (flu adj3 immuni$).tw.
5. (influenza adj3 vaccin$).tw.
6. flumist.tw.
7. (laiv adj2 vaccin*).tw.
8. (caiv-t adj2 vaccin*).tw.
9. or/1-8
10. exp Cardiovascular Diseases/
11. cardio*.tw.
12. cardia*.tw.
13. heart*.tw.
14. coronary*.tw.
15. angina*.tw.
16. ventric*.tw.
17. myocard*.tw.
18. pericard*.tw.
19. isch?em*.tw.
20. emboli*.tw.
21. arrhythmi*.tw.
22. thrombo*.tw.
23. atrial fibrillat*.tw.
24. tachycardi*.tw.
25. endocardi*.tw.
26. (sick adj sinus).tw.
27. exp Stroke/
28. (stroke or stokes).tw.
29. cerebrovasc*.tw.
30. cerebral vascular.tw.
31. apoplexy.tw.
32. (brain adj2 accident*).tw.
33. ((brain* or cerebral or lacunar) adj2 infarct*).tw.
34. exp Hypertension/

35. hypertensi*.tw.
36. peripheral arter* disease*.tw.
37. ((high or increased or elevated) adj2 blood pressure).tw.
38. exp Hyperlipidemias/
39. hyperlipid*.tw.
40. hyperlip?emia*.tw.
41. hypercholesterol*.tw.
42. hypercholester?emia*.tw.
43. hyperlipoprotein?emia*.tw.
44. hypertriglycerid?emia*.tw.
45. exp Arteriosclerosis/
46. exp Cholesterol/
47. cholesterol.tw.
48. "coronary risk factor* ".tw.
49. Blood Pressure/
50. blood pressure.tw.
51. or/10-50
52. 9 and 51
53. randomized controlled trial.pt.
54. controlled clinical trial.pt.
55. randomized.ab.
56. placebo.ab.
57. drug therapy.fs.
58. randomly.ab.
59. trial.ab.
60. groups.ab.
61. 53 or 54 or 55 or 56 or 57 or 58 or 59 or 60
62. exp animals/ not humans.sh.
63. 61 not 62
64. 52 and 63

**EMBASE Ovid**

1. influenza vaccine/
2. (influenza$ adj3 immuni$).tw.
3. (flu adj3 vaccin$).tw.
4. (flu adj3 immuni$).tw.
5. (influenza adj3 vaccin$).tw.
6. flumist.tw.
7. (laiv adj2 vaccin*).tw.
8. (caiv-t adj2 vaccin*).tw.
9. or/1-8
10. exp cardiovascular disease/
11. cardio*.tw.
12. cardia*.tw.
13. heart*.tw.

14. coronary*.tw.
15. angina*.tw.
16. ventric*.tw.
17. myocard*.tw.
18. pericard*.tw.
19. isch?em*.tw.
20. emboli*.tw.
21. arrhythmi*.tw.
22. thrombo*.tw.
23. atrial fibrillat*.tw.
24. tachycardi*.tw.
25. endocardi*.tw.
26. (sick adj sinus).tw.
27. exp cerebrovascular disease/
28. (stroke or stokes).tw.
29. cerebrovasc*.tw.
30. cerebral vascular.tw.
31. apoplexy.tw.
32. (brain adj2 accident*).tw.
33. ((brain* or cerebral or lacunar) adj2 infarct*).tw.
34. exp hypertension/
35. hypertensi*.tw.
36. peripheral arter* disease*.tw.
37. ((high or increased or elevated) adj2 blood pressure).tw.
38. exp hyperlipidemia/
39. hyperlipid*.tw.
40. hyperlip?emia*.tw.
41. hypercholesterol*.tw.
42. hypercholester?emia*.tw.
43. hyperlipoprotein?emia*.tw.
44. hypertriglycerid?emia*.tw.
45. exp Arteriosclerosis/
46. exp Cholesterol/
47. cholesterol.tw.
48. "coronary risk factor*".tw.
49. Blood Pressure/
50. blood pressure.tw.
51. or/10-50
52. 9 and 51
53. random$.tw.
54. factorial$.tw.
55. crossover$.tw.
56. cross over$.tw.
57. cross-over$.tw.
58. placebo$.tw.
59. (doubl$ adj blind$).tw.

60. (singl$ adj blind$).tw.
61. assign$.tw.
62. allocat$.tw.
63. volunteer$.tw.
64. crossover procedure/
65. double blind procedure/
66. randomized controlled trial/
67. single blind procedure/
68. 53 or 54 or 55 or 56 or 57 or 58 or 59 or 60 or 61 or 62 or 63 or 64 or 65 or 66 or 67
69. (animal/ or nonhuman/) not human/
70. 68 not 69
71. 52 and 70

**Web of Science**

#12 #11 AND #10
#11 TS=(random* or blind* or allocat* or assign* or trial* or placebo* or crossover* or cross-over*)
#10 #9 AND #1
#9 #8 OR #7 OR #6 OR #5 OR #4 OR #3 OR #2
#8 TS=(hyperlipid* OR hyperlip?emia* OR hypercholesterol* OR hypercholester?emia* OR hyperlipoprotein?emia* OR hypertriglycerid?emia*)
#7 TS=("high blood pressure")
#6 TS=(hypertensi* OR "peripheral arter* disease*")
#5 TS=(stroke OR stokes OR cerebrovasc* OR cerebral OR apoplexy OR (brain SAME accident*) OR (brain SAME infarct*))
#4 TS=("atrial fibrillat*" OR tachycardi* OR endocardi*)
#3 TS=(pericard* OR isch?em* OR emboli* OR arrhythmi* OR thrombo*)
#2 TS=(cardio* OR cardia* OR heart* OR coronary* OR angina* OR ventric* OR myocard*)
#1 TS=(((influenza* OR flu OR laiv OR caiv-t) NEAR/3 (immuni* OR vaccin*)) OR flumist)

**Limited PubMed search 20 February 2015**

((vaccine* OR vaccinat*) AND (influenza OR flu) AND (cardiovascular OR heart OR coronary OR stroke)) in PubMed Clinical Queries

Similar (adapted) search in www.controlled-trials.com and www.clinicaltrials.gov

### Appendix 2. Search strategies 2008 - coronary heart disease

**CENTRAL (2007, Issue 4)**

#1 INFLUENZA VACCINE
#2 (influenza* near vaccin*)
#3 (influenza* near immuni*)
#4 (flu near immuni*)

#5 (flu near vaccin*)
#6 (#1 or #2 or #3 or #4 or #5)
#7 CARDIOVASCULAR DISEASES
#8 heart
#9 coronary
#10 cardiac
#11 myocardial
#12 cardiovascular
#13 angina
#14 (#7 or #8 or #9 or #10 or #11 or #12 or #13)
#15 (#6 and #14)

**MEDLINE (to January 2008)**

1 Influenza Vaccine/
2 (influenza$ adj3 immuni$).tw.
3 (flu adj3 vaccin$).tw.
4 (flu adj3 immuni$).tw.
5 (influenza adj3 vaccin$).tw.
6 or/1-5
7 exp cardiovascular diseases/
8 myocardial.tw.
9 angina.tw.
10 coronary.tw.
11 heart.tw.
12 cardiac.tw.
13 cardiovascular.tw
14 or/7-13
15 6 and 14
16 randomized controlled trial.pt.
17 controlled clinical trial.pt.
18 Randomized controlled trials/
19 random allocation/
20 double blind method/
21 single-blind method/
22 or/16-21
23 exp animal/ not humans/
24 22 not 23
25 clinical trial.pt.
26 exp Clinical trials/
27 (clin$ adj25 trial$).ti,ab.
28 ((singl$ or doubl$ or trebl$ or tripl$) adj (blind$ or mask$)).ti,ab.
29 placebos/
30 placebo$.ti,ab.
31 random$.ti,ab.
32 research design/

33 or/25-32
34 33 not 23
35 34 not 24
36 comparative study.pt.
37 exp evaluation studies/
38 follow up studies/
39 prospective studies/
40 (control$ or prospectiv$ or volunteer$).ti,ab.
41 or/36-40
42 41 not 23
43 42 not (24 or 35)
44 24 or 35 or 43
45 15 and 44
46 limit 45 to yr="2005 - 2008"

**EMBASE (to January 2008)**

1 Influenza Vaccine/
2 (influenza$ adj3 immuni$).tw.
3 (flu adj3 vaccin$).tw.
4 (flu adj3 immuni$).tw.
5 (influenza adj3 vaccin$).tw.
6 or/1-5
7 exp cardiovascular disease/
8 myocardial.tw.
9 angina.tw.
10 coronary.tw.
11 heart.tw.
12 cardiac.tw.
13 cardiovascular.tw.
14 or/7-13
15 6 and 14
16 clinical trial/
17 random$.tw.
18 randomized controlled trial/
19 trial$.tw.
20 follow-up.tw.
21 double blind procedure/
22 placebo$.tw.
23 placebo/
24 factorial$.ti,ab.
25 (crossover$ or cross-over$).ti,ab.
26 (double$ adj blind$).ti,ab.
27 (singl$ adj blind$).ti,ab.
28 assign$.ti,ab.
29 allocat$.ti,ab.

30 volunteer$.ti,ab.
31 Crossover Procedure/
32 Single Blind Procedure/
33 or/16-32
34 15 and 33
35 limit 34 to yr="2005 - 2008"
